# Intervention strategies against COVID-19 and their estimated impact on Swedish healthcare capacity

**DOI:** 10.1101/2020.04.11.20062133

**Authors:** Jasmine M Gardner, Lander Willem, Wouter Van Der Wijngaart, Shina Caroline Lynn Kamerlin, Nele Brusselaers, Peter Kasson

## Abstract

**Objectives:** During March 2020, the COVID-19 pandemic has rapidly spread globally, and non-pharmaceutical interventions are being used to reduce both the load on the healthcare system as well as overall mortality.

**Design:** Individual-based transmission modelling using Swedish demographic and Geographical Information System data and conservative COVID-19 epidemiological parameters.

**Setting:** Sweden

**Participants:** A model to simulate all 10.09 million Swedish residents.

**Interventions:** 5 different non-pharmaceutical public-health interventions including the mitigation strategy of the Swedish government as of 10 April; isolation of the entire household of confirmed cases; closure of schools and non-essential businesses with or without strict social distancing; and strict social distancing with closure of schools and non-essential businesses.

**Main outcome measures:** Estimated acute care and intensive care hospitalisations, COVID-19 attributable deaths, and infections among healthcare workers from 10 April until 29 June.

**Findings:** Our model for Sweden shows that, under conservative epidemiological parameter estimates, the current Swedish public-health strategy will result in a peak intensive-care load in May that exceeds pre-pandemic capacity by over 40-fold, with a median mortality of 96,000 (95% CI 52,000 to 183,000). The most stringent public-health measures examined are predicted to reduce mortality by approximately three-fold. Intensive-care load at the peak could be reduced by over two-fold with a shorter period at peak pandemic capacity.

**Conclusions:** Our results predict that, under conservative epidemiological parameter estimates, current measures in Sweden will result in at least 40-fold over-subscription of pre-pandemic Swedish intensive care capacity, with 15.8 percent of Swedish healthcare workers unable to work at the pandemic peak. Modifications to ICU admission criteria from international norms would further increase mortality.

**What is already known?:**

- The COVID-19 pandemic has spread rapidly in Europe and globally since March 2020.
- Mitigation and suppression methods have been suggested to slow down or halt the spread of the COVID-19 pandemic. Most European countries have enacted strict suppression measures including lockdown, school closures, enforced social distancing; while Sweden has chosen a different strategy of milder mitigation as of today (10 April 2020).
- Different national policy decisions have been justified by socio-geographic differences among countries. Such differences as well as the tempo and stringency of public-health interventions are likely to affect the impact on each country’s mortality and healthcare system.

**What this study adds?:**

- Individual-based modelling of COVID-19 spread using Swedish demographics and conservative epidemiological assumptions indicates that the peak of the number of hospitalised patients with COVID-19 can be expected in early May under the current strategy, shifted earlier and attenuated with more stringent public health measures.
- Healthcare needs are expected to substantially exceed pre-pandemic capacity even if the most aggressive interventions considered were implemented in the coming weeks. In particular the need for intensive care unit beds will be at least 40-fold greater than the pre-pandemic capacity if the current strategy is maintained, and at least 10-fold greater if strategies approximating the most stringent in Europe are introduced by 10 April.
- Our model predicts that, using median infection-fatality-rate estimates, at least 96,000 deaths would occur by 1 July without mitigation. Current policies reduce this number by approximately 15%, while even more aggressive social distancing measures, such as adding household isolation or mandated social distancing can reduce this number by more than 50%.

## Introduction

COVID-19 has been spreading rapidly around the globe with a substantial effect on global morbidity, mortality and healthcare utilization. It was declared a pandemic by the World Health Organisation on 11 March 2020, at which time almost 1000 deaths were reported in Europe with confirmed community transmission in multiple European countries.^1^ In addition to marshalling therapeutic options, countries have adopted a range of non-pharmaceutical public health measures to reduce transmission. These have been broadly characterized as *suppressive* approaches, which aim to stop transmission through a mix of targeted and untargeted social measures, and *mitigation* approaches, which aim to slow the spread and shield vulnerable populations without ultimately interrupting transmission.^2 3^ Countries such as China and Singapore give examples of rapidly introduced suppressive strategies, while Sweden is the most prominent example of mitigation—limiting the extent of social distancing and economically disruptive interventions while still aiming to slow the spread sufficiently to allow for an effective medical response.^3 4^ Studying the Swedish strategy and its health implications thus yields important lessons for global public health policy.

COVID-19 is spreading globally because of a lack of prior immunity combined with relatively high infectiousness.^5 6 7^ Age-adjusted hospitalization rates range between 1% and 70% of reported cases in the United States^8^ and between 0.04% to 18.4% in China.^9^ This means that even with a substantial proportion of asymptomatic cases, the total number of people requiring hospitalization will exceed the total number of beds available in many countries.^10^ The ability to provide care thus depends on the tempo of pandemic spread. Based on the experiences from China, Italy and Spain, the critically limiting resource in reducing COVID-19 related mortality is the capacity of intensive care with mechanical ventilation – since many COVID-19 pneumonia patients die from severe respiratory diseases including acute respiratory distress syndrome (ARDS).^11 12^ Overall mortality for the outbreak in China was estimated to be 1.4% of the confirmed cases (0.7% including unconfirmed cases),^9^ yet figures up to 5.7% were reported for the Hubei province, which has been hit hardest.^13^

Socio-geographic differences between countries affect the speed and impact of the pandemic—these include population density, household structure, social contact patterns, and governmental measures implemented to hinder or stop the spread of the virus. While the pandemic is still ongoing and exact statistics are not available, numerical modelling provides timely guidance for healthcare needs given multiple scenarios, as it permits predictions based on available epidemiological and demographic data.^14^ Predictive models for COVID-19 have been used to assess the impact of the pandemic and guide national policy.^2 3 15-19^ While *aggregate* population models yield the basic principles of epidemic spread, *discrete* models that take into account geographic and demographic structure permit more specific assessment of different interventions in national contexts. Individual-based discrete models - also called agent-based models, since every individual of a geographic region is modelled by an agent - provide a flexible platform to analyse the propagation of emerging infectious diseases and the effect of social distancing and similar behavioural changes.^20^ In these individual-based models, the behaviour and health state of each individual in a country is modelled, enabling explicit representation of social-contact patterns between individuals in the household, school, workplace and community. These models similarly permit explicit representation of age- and location-specific social distancing such as school closure, increased telework, or geographic quarantine measures.

Because of Sweden’s unique strategy of not imposing stringent suppression strategies (until at least 10 April 2020), we used an individual-based model parameterized on Swedish demographics to assess the anticipated spread of COVID-19. We report the predicted outcomes of the current mitigation strategy versus Swedish healthcare capacity and assess alternative approaches.

## Methods

### Pre-pandemic setting and early phases of pandemic in Sweden

Sweden is the largest Northern European country with a population of 10 million individuals, a relatively low population density of 25 individuals/km^2^ and 3 large metropolitan areas: Stockholm, Gothenburg and Malmö.^21^ Sweden has the 8^th^ highest Human Development Index (0.937), with an average life expectancy at birth of 83 years.^21^ Based on data collected on European level, 204 “curative care” hospital beds were available per 100,000 inhabitants in Sweden (2017);^22^ and 5.8/100,000 intensive care unit (ICU) beds (2014) (European average respectively 372 and 11.5/100,000 inhabitants).^23 24^ This constitutes the lowest number of reported hospital beds and the second lowest number of ICU beds per capita in Europe.^22 24^ Most recent figures reported to the World Health Organisation in 2019 showed a 78% bed occupancy for acute care hospitals in Sweden (based on 1996 data, range 24-90%).^25^

The first case and first death of COVID-19 were reported on 31 January 2020 in Jönköping^26^ and 11 March 2020 in Stockholm, respectively, with the latter being a case of community transmission.^27 28^ The 10th death was recorded by 18 March, and the first 100 by 27 March.^29^ From 12 March, testing in Stockholm was only used for hospitalised patients from high risk groups and symptomatic health care staff,^27^ although testing has increased in early April.

### Individual-based model for pandemic spread

We employed an individual agent-based model based on work by Ferguson et al.^2 30 31^ Individual-based models are increasingly used to model epidemic spread with explicit representation of demographic and spatial factors such as population distribution, workplace data, school data, and mobility.^20^

*Individual properties*. Individuals are randomly assigned an age based on Swedish demographic data,^21^ and they are also assigned a household. Household size is normally distributed around the average household size in Sweden in 2018, 2.2 people per household.^10^

The relatively small household size (39.8% of Swedish households were single occupancy in 2019^21^) has been informally cited as a potential difference in Swedish infectious disease dynamics from the rest of Europe. Households are placed on a lattice using high-resolution population data^32^ with at least one household per square km that has a population ≥ 1. Households have at least one adult over 20 years old acting as a head of household. Additional members are randomly distributed throughout the households in each population square until the population is completely allocated subject to the population density dataset. Households have a maximum of 7 members. Each household is additionally allocated to a city based on the closest city centre by distance and to a county based on city designation.

Each individual is placed in a school or workplace at a rate similar to the current participation in Sweden. Three types of schools and two types of workplaces are described with some individuals not participating in the workforce. Children aged less than 1 or elderly adults older than 75 do not participate in the school community or workforce. Children aged 1-5 are randomly placed in a pre-school program within their city. Children aged 1-3 participate at a 78% rate of participation and children aged 3-5 participate at a rate of 95% participation in accordance with participation in Sweden as of 2016.^33^

Students aged 6 to 15 are randomly placed into elementary schools in their city with a size of 220 students per school (the average size of Swedish primary schools).^33^ Students aged 15 to 22 are randomly placed into higher education institutes in their county with similar school size. Workforce participation follows Swedish demographic data: 77.3% for individuals aged 22-65 and 17.2% for individuals aged 65-75.^21^ Workplaces are typically small in Sweden with approximately 95% of workplaces with 1-9 employees.^21^ Employees in our simulation are allocated to workgroups of 15 people to account for the skewing effect of larger workplaces and inter-workplace interactions in smaller ones. Overall, 4.325% of the workforce is assumed to work in healthcare in the hospital, calculated by assuming that 25% of the 17.25% of civilian employees who work in healthcare work in hospitals.^10^ These workers are in contact with other employees of the hospital as well as the hospital and ICU patients. Each hospital has an average of 120 employees prior to the pandemic. Each county has at least 1 hospital. Hospitals have only 25% of the infection spread of other workplaces to account for hospital infection-control precautions—this approximation is discussed further below.

#### Initial infections

Infections are initially seeded randomly within each county. The number of initial infections per county is based on COVID-19 ICU admissions from March 17, 2020 to March 31, 2020,^34^ using a calculation that 2.95% of total cases result in hospitalization, of which 30% result in ICU admission.^2^ Infections are distributed among counties with weights equal to the percentage of cases present in that county as of March 25, 2020.^29^

#### Disease transmission

Transmission between individuals occurs through contact at each individual’s workplace or school, within their household, and in their communities. Infectiousness is thus an additive property dependent on contacts from household members, school/workplace members and community members with a probability based on household distances. Infection spread is modelled through stochastic simulation. At each time step, the probability that each individual will be infected is based on the contact risk with prior infected individuals with a probability of 1−exp(−λ_i_ΔT), where λ_i_ is the infectiousness parameter for individual i.

#### Transmissibility scaling factors

Transmissibility was calibrated against data for the period 21 March – 6 April to reproduce either the doubling time reported using pan-European data^35^ or the growth in reported Swedish deaths for that period.^27^ This yielded two distinct doubling rates: 3 days and 5 days, which were applied respectively to each of the model runs as transmission scaling factors.

#### Statistical sampling

The model was simulated with doubling times of 3 days (100 runs, doubling time from European data) and 5 days (10 runs, based on growth in Swedish deaths). 95% confidence intervals are reported representing statistical sampling error across these runs in all cases except for total deaths, where the 95% confidence intervals for age-adjusted infection fatality rates in China^9^ are propagated through the model.

Further details on the transmission dynamics are described in **Supplement 1**.

### Interventions

Public-health interventions are introduced to reduce the spread of transmission. These interventions can be required for all individuals, i.e. for school closures, or have a compliance rate associated to account for individuals who do not follow the mandate. The following interventions were tested and analysed.

1) Current Swedish policy: This intervention emulates Swedish government policy through at least 10 April 2020. In this intervention, high schools and universities are closed, people aged 70 and above are advised to practice social distancing, and symptomatic people are advised to practice home quarantine. All students in high school or college (aged 15-22 years old) do not attend school as implemented by the Swedish authorities, effective 13 March 2020. Their school transmission rate is eliminated. For these students, community transmission is increased by 25% and household transmission is increased by 50%. Individuals over 70 years of age reduce workplace and community contacts by 75%. Symptomatic people home quarantine after 1 day of symptoms by removing themselves from the workplace, reducing community transmission by 75%, and maintaining household transmission with a 90% compliance rate. All additional interventions are overlaid on this policy.

2) Case isolation of entire households: In addition to intervention 1, all individuals in a household with a symptomatic person are advised to self-quarantine. During household quarantine, community transmission is reduced by 75%, workplace transmission is removed, and household transmission is increased by 50%. Compliance is randomly determined at time of infection with an overall compliance rate of 70%. If the non-infected household member becomes infected, they practice symptomatic self-quarantine without increased household transmission at a 90% compliance rate.

4) Closure of all schools: In addition to intervention 1, all schools including preschools, elementary schools, and high schools/colleges will be closed. All school interactions are removed, interactions between children and the household increase by 50% and interactions between children and the community increase by 25%.

5) Closure of schools and non-essential businesses without social distancing: In addition to intervention 1, all schools and non-essential businesses are closed. Social distancing is not advised. School transmission is removed, workplace transmission is reduced by 75%. Household interactions increase by 50% and community interactions increase by 50%. This essentially emulates a public holiday.

6) Social distancing with closure of schools and non-essential businesses: In addition to intervention 1, all schools and non-essential businesses are closed, and social distancing is advised for the entire population. School transmission is removed, workplace transmission is reduced by 75%. Household interactions increase by 50% and community interactions decrease by 75%. Social distancing occurs at a 90% compliance rate, indicating strong reduction of all social contacts.

### Implementation and data sharing

The code implementing this model and the interventions tested will be made freely available on https://github.com/kassonlab/covid19-epi. Simulations were run on resources provided by the Swedish National Infrastructure for Computing (SNIC), at the PDC Centre for High Performance Computing (PDC, Stockholm), the High Performance Computing Centre North (HPC2N, Umeå), the Uppsala Multidisciplinary Centre for Advanced Computational Science (UPPMAX, Uppsala), and the National Supercomputer Centre (NSC, Linköping), as well as local resources at the University of Virginia. Results files are available on Zenodo (DOI:10.5281/zenodo.3748120).

### Patient and public involvement

Since the COVID-19 pandemic is still ongoing and rapidly spreading in Sweden, the results will be of high importance to the Swedish public; and will be disseminated broadly to the general public and the Swedish authorities. All authors are affiliated with Swedish or European universities, and are therefore highly involved from a personal perspective, as is the entire Swedish (and European) population, as the policies implemented by individual European countries will have effects on the whole continent, and thus be felt at both the European and global level.

### Ethics approval

Since the study was based on publicly available data, ethics approval was not required.

## Results

Because RT-PCR testing is not comprehensively performed, models were initialized using numbers of infected persons back-calculated from ICU admissions. Parameters were validated against the reported COVID-19 death rates in Sweden from 21 March to 6 April. All models were run approximating the social-distancing measures implemented by the Swedish public health authorities from 21 March to 10 April, and each public-health intervention evaluated was separately implemented on 10 April.

### Current strategy (without strict suppression measures)

Strikingly, models of the current public-health interventions predicted a rapid rise in individuals infected with SARS CoV-2, with 50% of susceptible individuals becoming infected within 30 days since 21 March in models parameterized to a doubling time of 3 days and 37 days in models parameterized to a doubling time of 5 days. This predicts a relatively narrow window for public-health interventions and a relatively high flux of patients through the healthcare system. Time-curves for predicted numbers of infected patients, hospitalized patients, ICU-patients and deaths are plotted in **Figures 1-4**. Taking Sweden as a whole, without further intervention, and presuming “standard” ICU admission criteria without restriction to ICU care (new ICU admission guidelines for COVID-19 have been communicated March 17, 2020 for Stockholm region^36^), the model predicts a required ICU capacity of at least 20,000 beds if continuing with current recommendations, with a peak in the first half of May.

**Figure 1.**
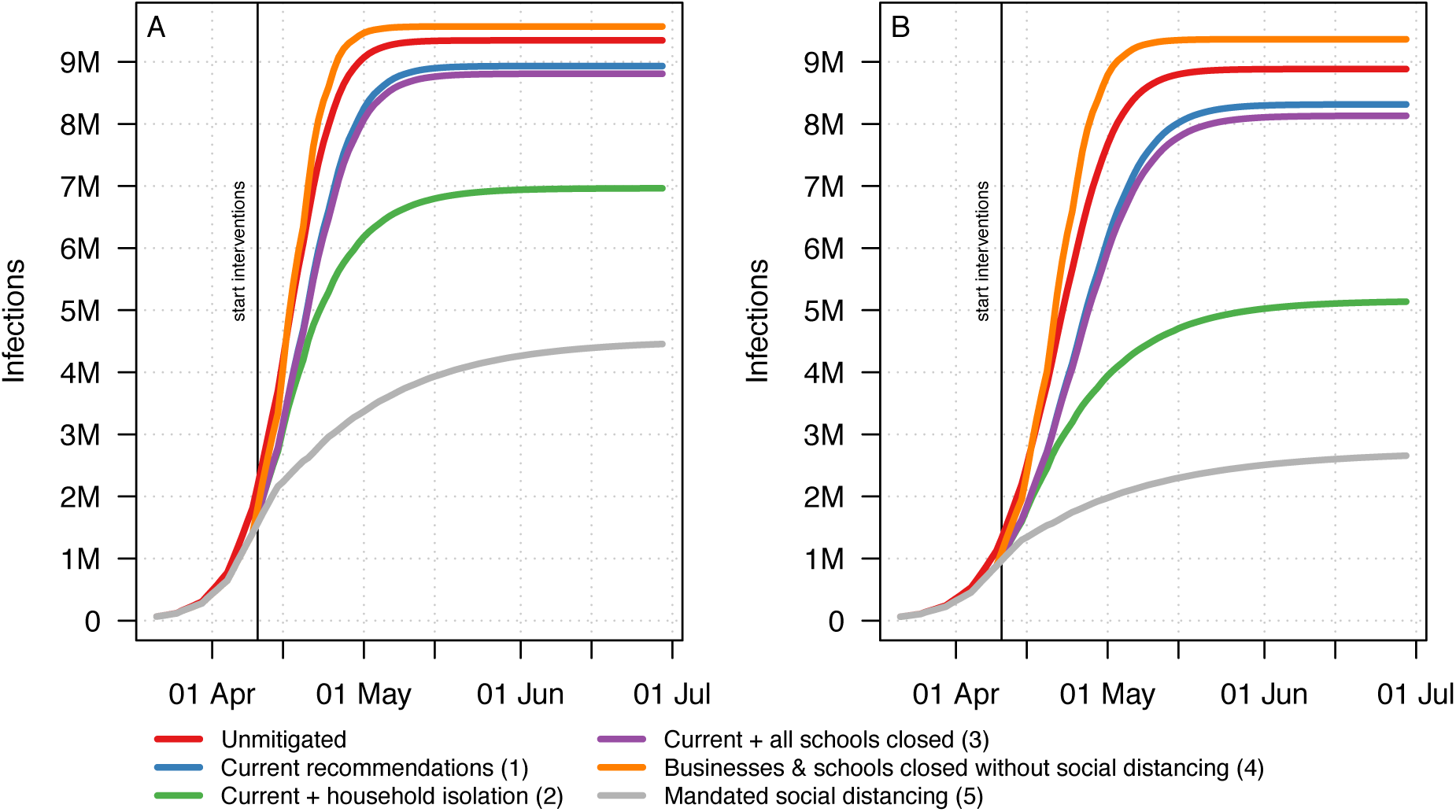
Estimated number of COVID-19 infections. Infections are plotted for each public health intervention strategy over time, using models parameterized to doubling times of (A) 3 or (B) 5 days. For a description of each intervention modelled, see the Methods.

**Figure 2.**
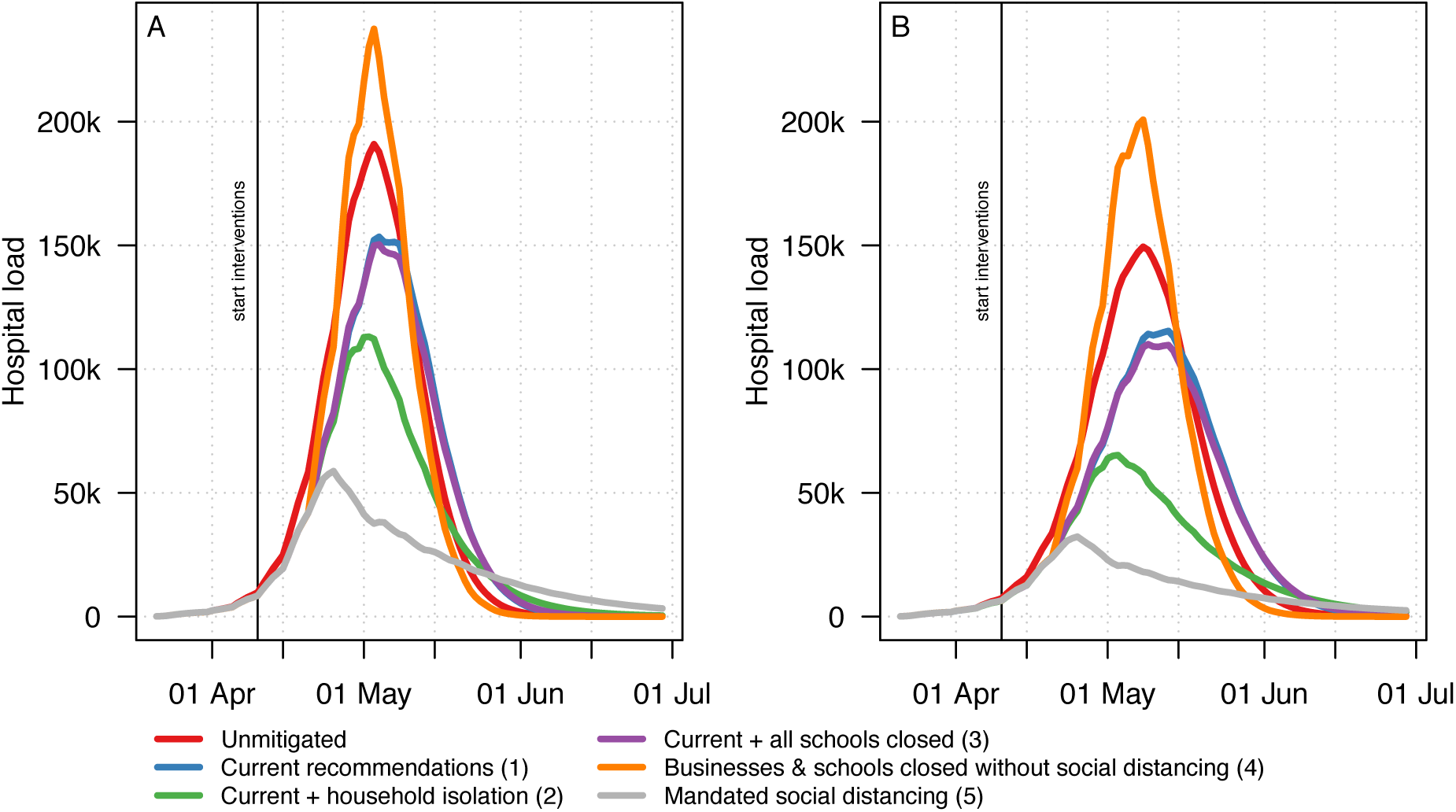
Estimated number of COVID-19 related hospitalisations. Hospitalizations are plotted for each public health intervention strategy over time, using models parameterized to doubling times of (A) 3 or (B) 5 days. For a description of each intervention modelled, see the Methods.

### Effect of social distancing interventions

Using the strictest intervention evaluated (mandated social distancing), the number of infections and hospitalisations (**Figures 1** and **2**) would each drop by at least twofold compared to the current strategy.

Since lack of mechanical ventilation will likely result in death for patients who would otherwise need it, it is worthwhile to consider alternative public-health intervention strategies to reduce the load on the healthcare system. The most stringent control measures considered resulted in at least a 50% reduction in the need for ICU capacity, bringing it to within an order of magnitude of pre-pandemic levels, with decline already beginning in late April. The predicted ICU capacity requirements associated with each public-health policy choice modelled are plotted in **Figure 3**. Because of Sweden’s low number of ICU beds per capita^22 24^, it likely had a high ICU-bed occupancy before the pandemic, so not all ventilators and staff will be available for COVID-19 patients even if all elective procedures are halted.

**Figure 3.**
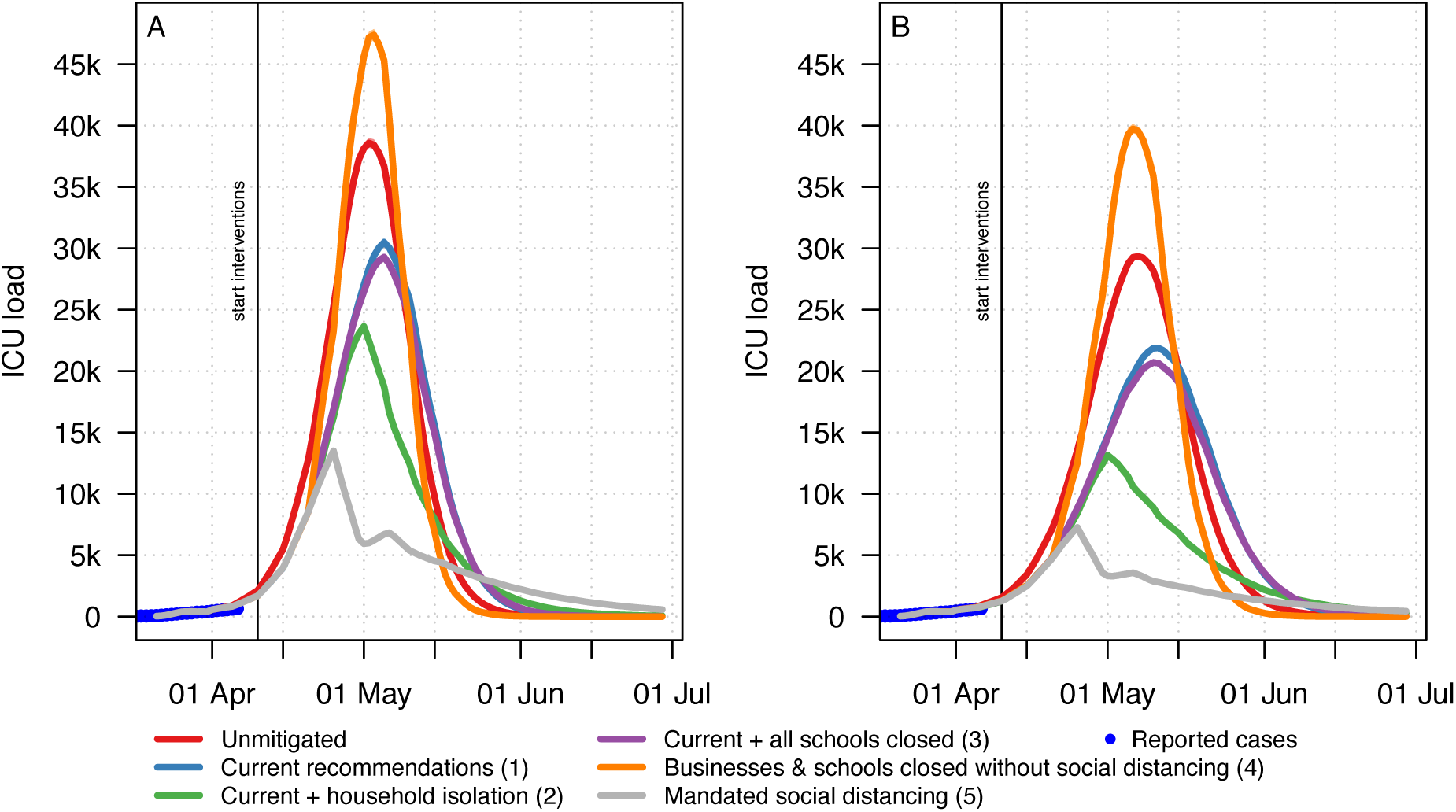
Estimated number of COVID-19 related intensive care unit hospitalisations. ICU census is plotted for each public health intervention strategy over time, using models parameterized to doubling times of (A) 3 or (B) 5 days. For a description of each intervention modelled, see the Methods.

For the total accumulated number of deaths (**Figure 4**), without mitigation, we predict at least 100,000 deaths by 1 July in both models. The current recommendations reduce this number by less than 20%. Aggressive social distancing measures, such as adding household isolation or mandated social distancing can reduce this number by at least 50%.

**Figure 4.**
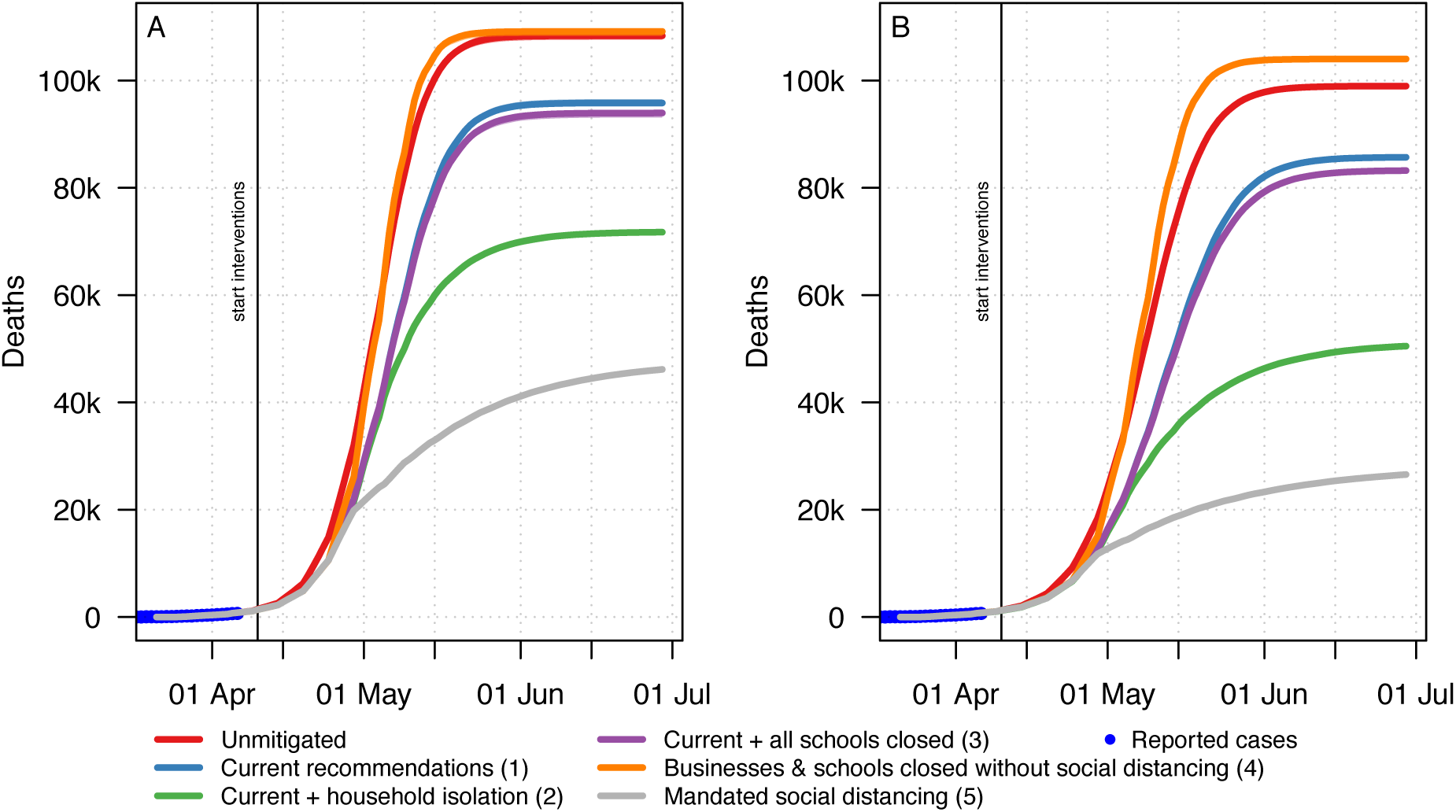
Estimated number of COVID-19 related deaths. Deaths are plotted for each public health intervention strategy over time, using models parameterized to doubling times of (A) 3 or (B) 5 days. For a description of each intervention modelled, see the Methods. Confidence intervals based on infection-fatality rates are listed in the text.

### Healthcare workers

Finally, we also considered infections among healthcare workers. Healthcare workers can become infected via three mechanisms: “community” exposure in their households or outside the hospital, transmission among healthcare staff during breaks and other times when personal protective equipment (PPE) is not worn, or occupational exposure to infected patients (either identified as such or not). Given sufficient PPE, occupational exposure should be minimal, and this has been shown in other well-resourced settings. However, at high patient load, PPE has been limiting in a large number of countries, and the Swedish PPE recommendations have recently been reduced,^37 38^ potentially in response to PPE shortages. We approximate a 75% reduction in transmissibility within the hospital in order to capture the aggregate effect of PPE usage and non-PPE encounters. There are substantial uncertainties in this estimate--healthcare workers without PPE are at increased risk of infection, so in the absence of universal PPE protocols, unidentified COVID-19 patients pose a higher probability of transmission in the healthcare context than outside. Transmission among healthcare workers through contact with colleagues is presumed to be similar to other workplaces, and, while using appropriate PPE, transmission should be small. Under this gross approximation, we predict substantial numbers of healthcare workers to become infected under the current public-health policies. In particular, 90% of healthcare workers could become infected with 1.5% of total healthcare workers in need of hospitalisation at the peak and 15.8% unable to work due to severe infection, which could affect the healthcare capacity. Note that these estimates are based on the availability of rigorous PPE for healthcare workers; obviously without proper PPE these numbers will be higher. Under mandated social distancing measures, fewer healthcare workers would be infected and thus excluded from the workforce. This is plotted in **Figure 5**. We note that even when healthcare workers are infected at home, PPE is a critical consideration for the health of these workers—anecdotal evidence suggests that occupational exposure of healthcare workers can lead to poorer outcomes than community exposure.

**Figure 5.**
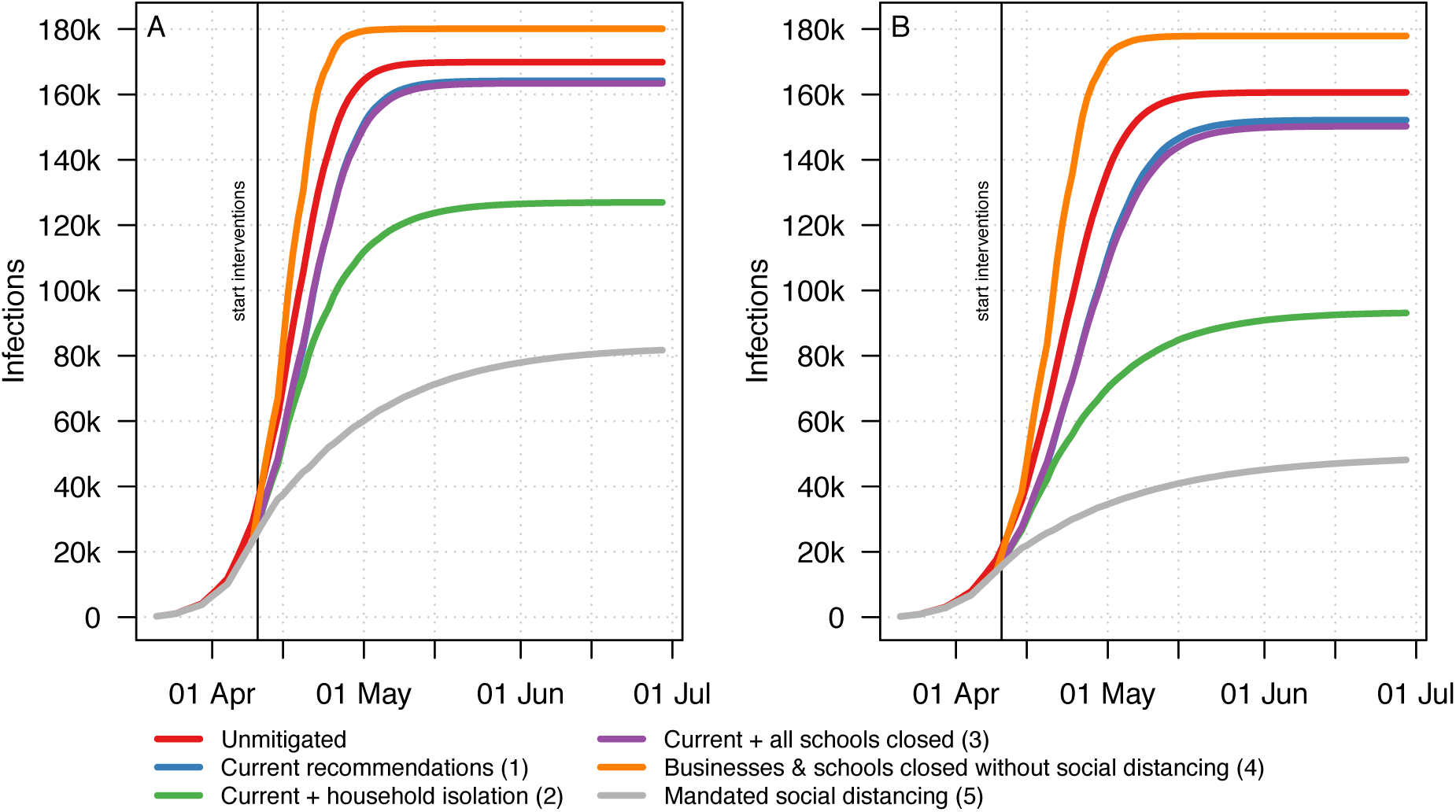
Estimated number of COVID-19 related healthcare worker infections. Healthcare worker infections are plotted for each public health intervention strategy over time, using models parameterized to doubling times of (A) 3 or (B) 5 days. For a description of each intervention modelled, see the Methods.

## Discussion

### Statement of principal findings

This individual-based modelling project predicts that with the current mitigation approach approximately 96,000 deaths (95% CI 52,000 to 183,000) can be expected before 1 July, 2020. At the peak period (early May), the need for ICU beds will be at least 40-fold higher than the pre-pandemic ICU-bed capacity, not considering ICU admissions for other conditions.

### Strengths and weaknesses of the study

The model predictions presented here utilize the best international understanding of the parameters of COVID-19 infectivity, clinical course, and transmissibility. Key parameters were further estimated from the Swedish data that we believe most robust to potential testing bias: ICU admissions and deaths. Based on this, the model makes striking predictions for the course of infection in Sweden over the coming weeks, based on Swedish government policy through 10 April 2020. In addition to estimating the results of public-health policy decisions on healthcare capacity and mortality, these predictions also provide a way to test current knowledge of the disease. Should either those reported data or current global understanding of COVID-19 biology include substantial errors, those will become evident as a divergence between model predictions and Sweden’s public health situation. In addition, residential care for the elderly is not accounted for explicitly in the models; this may lead to incorrect estimation of mortality in the older age groups.

#### Strengths and weaknesses in relation to other studies, discussing important differences in results

Sweden is selecting a public-health strategy for COVID-19 different from that of most other industrialized countries,^20 39^ A number of epidemiological models have suggested that the current strategy could have dire consequences,^4 40^ but one persistent criticism has been whether those models take into account the social and demographic structure of Sweden. Here, we analyse COVID-19 spread in Sweden using an individual-based model^20^ that explicitly encodes the geographic and demographic structure of Sweden. We have used conservative estimates based on preliminary data from Sweden and other countries, and monitored the reported number of COVID-19 diagnoses, hospital and ICU admissions as well as deaths closely during the process of designing and running these prediction models. Testing has been restricted to high risk hospitalised cases and healthcare workers until early April, and only laboratory-confirmed cases and deaths were counted in the official statistics in accordance to the Infection Protection Act, and there is a delay in reporting.^27^

### Meaning of the study: possible explanations and implications for clinicians and policymakers

Our results predict that Sweden’s healthcare capacity (both for COVID-19 patients and for others) will be rapidly overwhelmed under the current strategy, both through the need to care for COVID-19 patients and because healthcare workers will themselves become ill and unable to work.^41-49^ Intensive care capacity is projected to become a particular bottleneck, as shown in other countries.^50-52^ For Sweden, this is in particular worrisome considering the relatively low availability of beds prior to the pandemic compared to other European countries^22 24^ and recent reports challenging Swedish disaster preparedness.^53^ We predict that aggressive suppressive measures can substantially reduce this healthcare capacity deficit but only if implemented in a timely manner. Whereas the predictive value of our model should become much clearer over the next two weeks, suppressive measures should be taken more immediately to have a substantial effect, especially for the Stockholm region. The projected mortality from COVID-19 may cause a Swedish all-cause mortality in 2020 exceeding that of 1918, which was the greatest absolute mortality in Sweden in the 20^th^ century. The COVID-19 pandemic and associated public-health measures will also impact non-COVID-19 related healthcare and mortality, as well the country’s economy. These factors are beyond the scope of the present project.

### Unanswered questions and future research

Due to the rapid spread of COVID-19, the window for such suppressive measures is relatively short. Our predictions are that if Sweden wishes to change course, prompt and decisive action would be required. The results of Sweden’s public-health policy will inform the world’s knowledge of COVID-19 and the efficacy of suppressive measures or lack thereof. We hope that such knowledge will not come at substantial cost of life.

## Data Availability

The code implementing this model and the interventions tested will be made freely available on https://github.com/kassonlab/covid19-epi. Results files are available on Zenodo (DOI:10.5281/zenodo.3748120).

https://doi.org/10.5281/zenodo.3748119

## Acknowledgements

LW received funding from the Research Foundation Flanders (1234620N) and from the Epipose project from the European Union’s SC1-PHE-CORONAVIRUS-2020 programme (101003688). SCLK and PK received funding from the Knut and Alice Wallenberg Foundation (2018.0140 and 2019.0431).

The simulations were enabled by resources provided by the Swedish National Infrastructure for Computing (SNIC 2020/5-176) at HPC2N in Umeå, UPPMAX in Uppsala, NSC in Linköping, and PDC in Stockholm, partially funded by the Swedish Research Council through grant agreement no. 2016-07213.

Åke SANDGREN (HPC2N) and Lars VIKLUND (HPC2N) are acknowledged for assistance concerning technical and implementational aspects, and in optimizing the performance of the code at the relevant centre-provided resources. Tove FALL (Uppsala University), Stefan ENGBLOM (Uppsala University), Nils FALL (Swedish University of Agricultural Sciences), Joacim ROCKLÖV (Umeå University), and Cecilia SÖDERBERG NAUCLÉR (Karolinska Institutet) are acknowledged for their important contributions to the design of the project.

## Supplements

Supplement 1: Supplementary methods

